# Post-COVID syndrome prevalence and risk factors in children and adolescents: A population-based serological study

**DOI:** 10.1101/2022.08.24.22279150

**Authors:** Roxane Dumont, Viviane Richard, Elsa Lorthe, Andrea Loizeau, Francesco Pennacchio, María-Eugenia Zaballa, Hélène Baysson, Mayssam Nehme, Anne Perrin, Arnaud G. L’Huillier, Laurent Kaiser, Rémy P. Barbe, Klara M. Posfay-Barbe, Silvia Stringhini, Idris Guessous, the SEROCoV-KIDS study group

## Abstract

**Objectives:** Post-COVID syndrome remain poorly studied in children and adolescents. In this study, we aimed to investigate the prevalence and risk factors of pediatric post-COVID in a population-based sample, stratifying by serological status.

**Study design:** We used data from the SEROCoV-KIDS cohort study (State of Geneva, Switzerland), which included children (aged 6 months to 17 years) selected from random samples drawn from state registries or who had a household member participating in a COVID-19 seroprevalence study conducted by our group. Children were tested for anti-SARS-CoV-2 N antibodies. Parents filled in a questionnaire on persistent symptoms in their children (lasting over 12 weeks) compatible with post-COVID syndrome.

**Results:** From December 1^st^, 2021 to February 16^th^, 2022, 1034 children were included, among whom 570 (55.1%) were seropositive. The sex- and age-adjusted prevalence of persistent symptoms among seropositive children was 9.1% (95%CI: 6.7;11.8) and 5.0% (95%CI: 3.0;7.1) among seronegatives, with an adjusted prevalence difference (ΔaPrev) of 4.1% (95%CI: 1.1;7.3). After stratification by age group, the prevalence was higher among adolescents aged 12-17 years (ΔaPrev=8.3%, 95%CI: 3.5;13.5) than among younger children (0.0%, 95%CI: −5.2;5.2 among 6-11 years old and 4.2%; 95%CI: −4.4;13.3 among 0-5 years old). The most frequently declared persistent symptoms among seropositives were smell loss, trouble concentrating and abdominal pain. Older age, having a chronic condition and lower socioeconomic conditions were identified as risk factors.

**Conclusion:** A significant proportion of seropositive children, particularly adolescents, experienced persistent symptoms. While there is a need for further investigation, growing evidence of pediatric post-COVID syndrome urges early screening and primary care management.

## Introduction

Evidence to date indicates that children, like adults, can experience post-COVID syndrome, also known as long COVID, with possibly major consequences on daily life (1). The scientific community agreed on its definition in the pediatric population in March 2022 (2): “Post-COVID-19 condition occurs in young people with a history of confirmed SARS-CoV-2 infection, with at least one persisting physical symptom for a minimum duration of 12 weeks after initial testing that impacts everyday functioning and cannot be explained by an alternative diagnosis”.

Many uncertainties remain regarding its prevalence, diagnosis, duration and treatment (3,4), partly due to clinical and methodological challenges (5). In a recent systematic review (3) (22 studies from 12 countries including 23,141 children and adolescents, of which 5 studies had population-based control groups), the prevalence of post-COVID symptoms (lasting more than three months) was 2 to 8% higher in the seropositive than in the control group, with a higher prevalence difference in adolescents. This prevalence was lower than suggested by others studies that did not use control groups (4), which may be due to several explanations. First, many children might experience long-lasting symptoms due to other viruses or medical events or due to the general stressful pandemic environment. Therefore, differentiating symptoms linked specifically to a SARS-CoV-2 infection from other diagnoses using a control group of non-infected persons is paramount to avoid overestimation of post-COVID syndrome prevalence. Second, most studies on post-COVID syndrome rely on samples of confirmed infection (RT-PCR and antigen tests) (3,6) hence excluding asymptomatic cases and underestimating the proportion of infected children and adolescents as this population was not systematically tested. Serological data allow to precisely estimate the proportion of infected children and adolescents by including asymptomatic and mild cases. Accordingly, the difference of prevalence between seropositives and seronegatives can provide an accurate estimator of post-COVID syndrome prevalence and/or unexpected complications such as acute hepatitis in the general pediatric population. The most frequently declared pediatric post-COVID symptoms are fatigue, headache, shortness of breath, chronic cough and myalgia with a higher risk among girls and adolescents (4). Adults also develop similar symptoms, including persistent cough, fever, headache, chest pain, hair loss, loss of taste and smell, amongst many others (7).

In addition to the uncertainties previously mentioned, only very few studies have so far analysed risk factors of pediatric post-COVID syndrome. A large body of literature has shown the adverse effects of low socioeconomic conditions on several health outcomes (8). Similar mechanisms could be expected in the pediatric post-COVID population.

We aimed to assess the prevalence and risk factors of persistent pediatric symptoms lasting over 12 weeks stratified by age group and anti-SARS-CoV-2 serological status.

## Methods

### Study design and data collection

Data are drawn from the SEROCoV-KIDS study, an ongoing, longitudinal and prospective cohort study, which aims at monitoring and evaluating direct and indirect impacts of the COVID-19 pandemic on the health and development of children and adolescents.

Children and adolescents’ eligibility criteria included 1) being between 6 months and 17 years old 2) residing in the Canton of Geneva at the time of enrolment, and 3) either being newly selected from random samples obtained from state registries or having a household member already participating in a population-based COVID-19 seroprevalence study conducted by our group (9–11). These population-based samples were provided by the Swiss Federal Statistical Office (FSO).

At the baseline assessment, all children were invited to perform a serological test (by blood drawing) to measure anti-SARS-CoV-2 antibodies (anti-N). One of the parent or legal guardian (referent parent) was asked to fill out online questionnaires related to health and development for him/herself and for each of his/her children, on the Specchio-COVID19 secured digital platform (12).

The Geneva Cantonal Commission for Research Ethics approved this study (ID 2021-01973). Referent parents of participants, as well as adolescents aged 14 years or older, provided written informed consent. Younger children provided oral consent.

The baseline assessment consisted of a blood sample for serological testing and online questionnaires. Non-respondents received email reminders 4, 7 and 21 days after the first questionnaire invitation and up to two phone calls were made.

### Study population

For this analysis, we selected all children who were included from December 1^th^, 2021 until February 16^th^, 2022. Children for whom we failed to take a blood sample were excluded from this analysis.

### Symptoms evaluation

At baseline, parents were systematically asked if their child(ren) had suffered from symptoms lasting at least 4 weeks since the beginning of the pandemic, no matter their SARS-CoV-2 infectious status. The duration of symptoms was then classified as lasting 4 to 6 weeks, 6 to 8 weeks, 8 to 12 weeks or more than 12 weeks. Parents had to report the approximate date of the start of persistent symptoms. They could select symptoms from an exhaustive list of COVID-19-related symptoms based on a literature review at the time of the questionnaire design and revised by a group of expert physicians on post-COVID. The persistent symptoms were grouped into seven general categories: general, respiratory, gastrointestinal, cardiovascular, musculoskeletal, neurological and dermatological symptoms. The severity of the persistent symptoms was then evaluated for the persistent symptoms using the following question asked to the parent “how much did the symptoms affect the child’s daily life?” (On a scale ranging from 1: very low limitation to 10: strong limitation). Parents were not aware of their child(ren) serological test while answering the questionnaire.

### SARS-CoV-2 infection

Confirmed SARS-CoV-2 infection was defined from a positive SARS-CoV-2 test (RT-PCR or antigen tests) reported by parents, along with the date of the first positive test. Parent could also declare whether the child(ren) had experience an episode of acute symptoms that could be considered as a symptomatic SARS-CoV-2 infection (confirmed symptomatic infection). Serological tests were based on the semiquantitative commercially-available immunoassay Roche Elecsys anti-SARS-CoV-2 N, detecting total Ig (including IgG) against the nucleocapsid protein of the SARS-CoV-2 virus. Seropositivity was defined using the manufacturer’s cut-off of index ≥1.0)(13). The test has an in-house sensitivity of 99.8% (95% CrI, 99.4%-100%) and specificity of 99.1% (95% CI, 98.3%-99.7%) (Roche Diagnostics, Rotkreuz, Switzerland). The antibodies detected by this test (anti-N antibodies) are produced following an infection but not following vaccination with mRNA vaccines, the only ones approved to date for children in Switzerland.

### Other characteristics

Other variables were described, including sociodemographic characteristics such as age, sex, parental education and household financial status. Parental education (based on the referent parent) was categorized into three groups: primary education (compulsory schooling), secondary education (apprenticeship and high school) and tertiary education (universitary studies). Household financial status was defined as “average to poor” if the referent parent chose one of the following statements about their financial situation: “I have to be careful with my expenses and an unexpected event could put me into financial difficulty” or “I cannot cover my needs with my income and I need external support”. If the referent parent had selected the following statement “I am comfortably off, money is not a concern and it is easy for me to save” or “My income covers my expenses and covers any minor contingencies” about their household financial situation was defined as “good”. Referent parents were also asked if their child(ren) suffered from a chronic health condition, defined as a medical/physical condition diagnosed by a health professional that lasted (or was expected to last) longer than 6 months.

### Statistical analysis

We compared sociodemographic and health-related characteristics between children who tested positive for anti-SARS-CoV-2 N antibodies and children who tested negative, overall and stratified into three age groups (0 to 5, 6 to 11 and 12 to 17 years old) using Chi-squared and Student-t tests, as appropriate.

We used marginal prediction after logistic regression to estimate prevalence and prevalence difference, adjusting for age and sex. Mixed-effect Poisson regression with robust variance, based on the sandwich estimator (14), was used to estimate prevalence ratio (15) and correct for potential dependence between participants as some children were siblings. Participants with missing data in at least one of the covariates (n=9, 1%) were excluded from the models. Statistical significance was defined as a level of confidence of 95% and all analyses were performed with R (version 4.0.3).

## Results

### Descriptive results

Among 3060 households, 625 households participated in our study (participation rate of 20.4%) (Figure 1).

**Figure 1:**
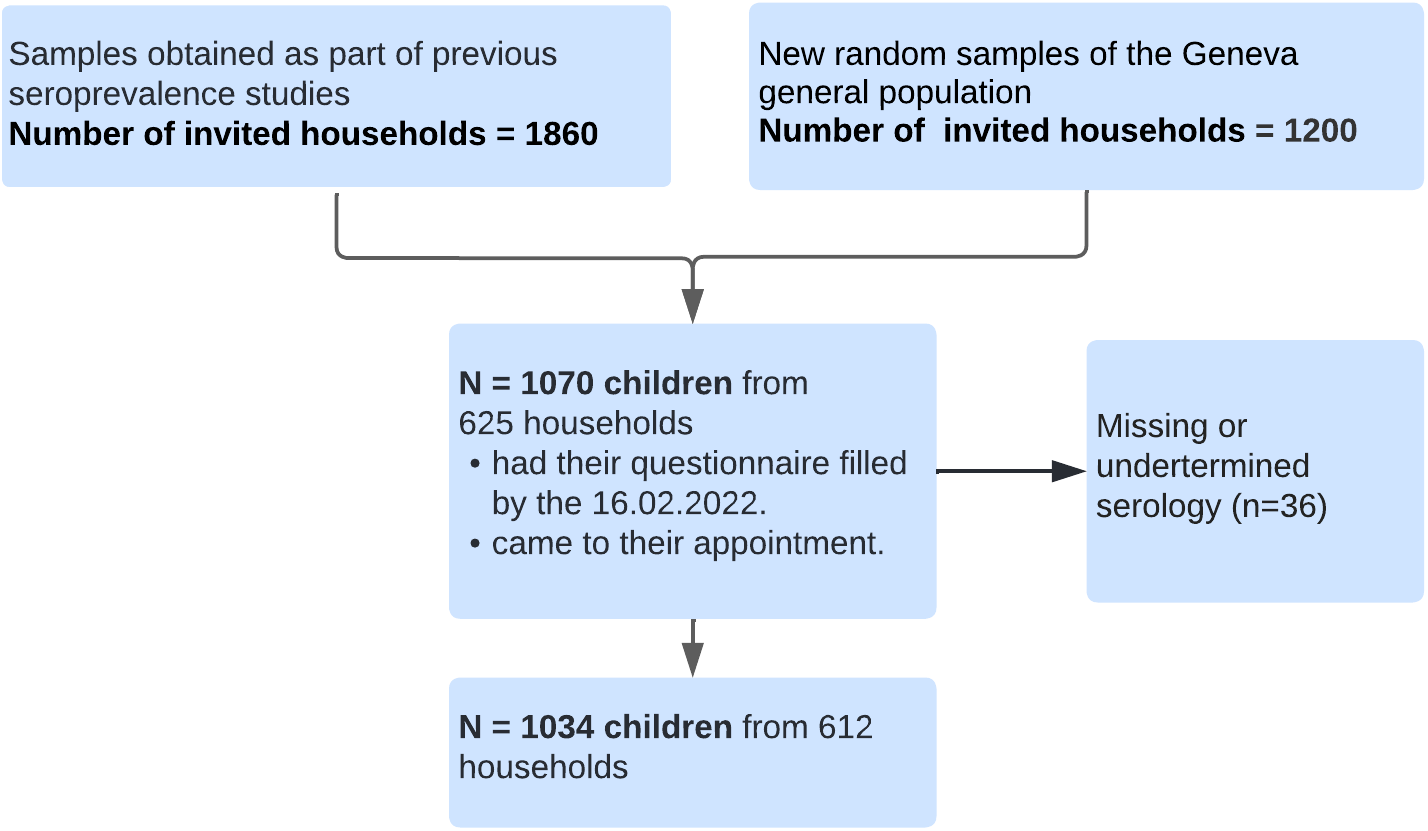
Flowchart

Our sample included 1034 children aged 6 months to 17 years from 612 households: 505 (49%) were girls and the mean age was 10.2 years [SD=4.2]). Overall, 785 (76%) referent parents had a tertiary education, 200 (19%) a secondary education and 42 (4%) had a primary education level (7 missing data). Among our participants, 150 (15%) were reported to live in a household with an average to poor financial situation; and 270 (26%) were reported to have a chronic medical condition (Table 1, S1). There were 570 (55%) children who tested positive for anti-SARS-CoV-2 N antibodies. Overall, we observed that 253 (24%) had a documented confirmed COVID-19 infection, and 198 (19%) declared an episode of acute symptoms that could be considered as a symptomatic SARS-CoV-2 infection. Among the 79 (8%) children who had experienced persistent COVID-19-related symptoms lasting over 12 weeks since the beginning of the pandemic, 54 (68%) were seropositive and 25 (32%) were seronegative.

**Table 1:**
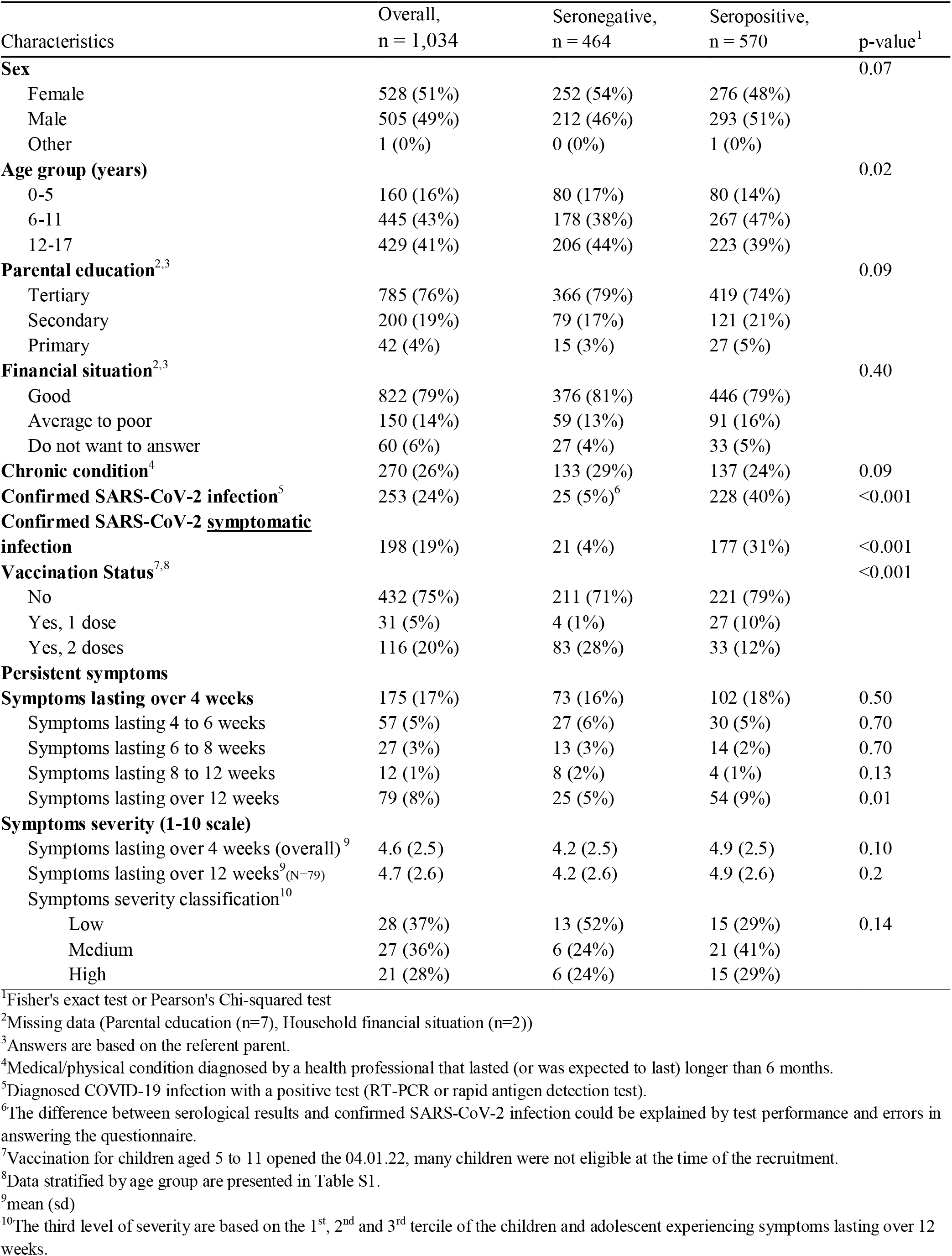
Descriptive statistics

Compared to seronegatives, seropositive children tended to experience more often symptoms such as smell loss, trouble concentrating, abdominal pain, fatigue, muscle pain, breathing difficulties and palpitations. In contrast, seronegative children were declared as having more anxiety, cough, dripping nose and dermatological symptoms (skin rash). Only smell and taste loss and palpitation were considered significantly more common in seropositive subjects (Figure 2).The severity of symptoms lasting over 12 weeks was on average slightly higher among seropositive children (mean [sd]=4.9 [2.5]) compared to seronegative subjects (mean [sd]=4.2 [2.5]), although not statistically significant (p-value=0.1) (Table 1). Among the 54 (9.5%) seropositive children who suffered from symptoms lasting over 12 weeks (Table S2), 31 (57%) were adolescents aged 12 to 17 years. A confirmed infection was documented in 30/54 (56%) and 26/54 (48%) reported acute symptoms at the time of infection.

**Figure 2:**
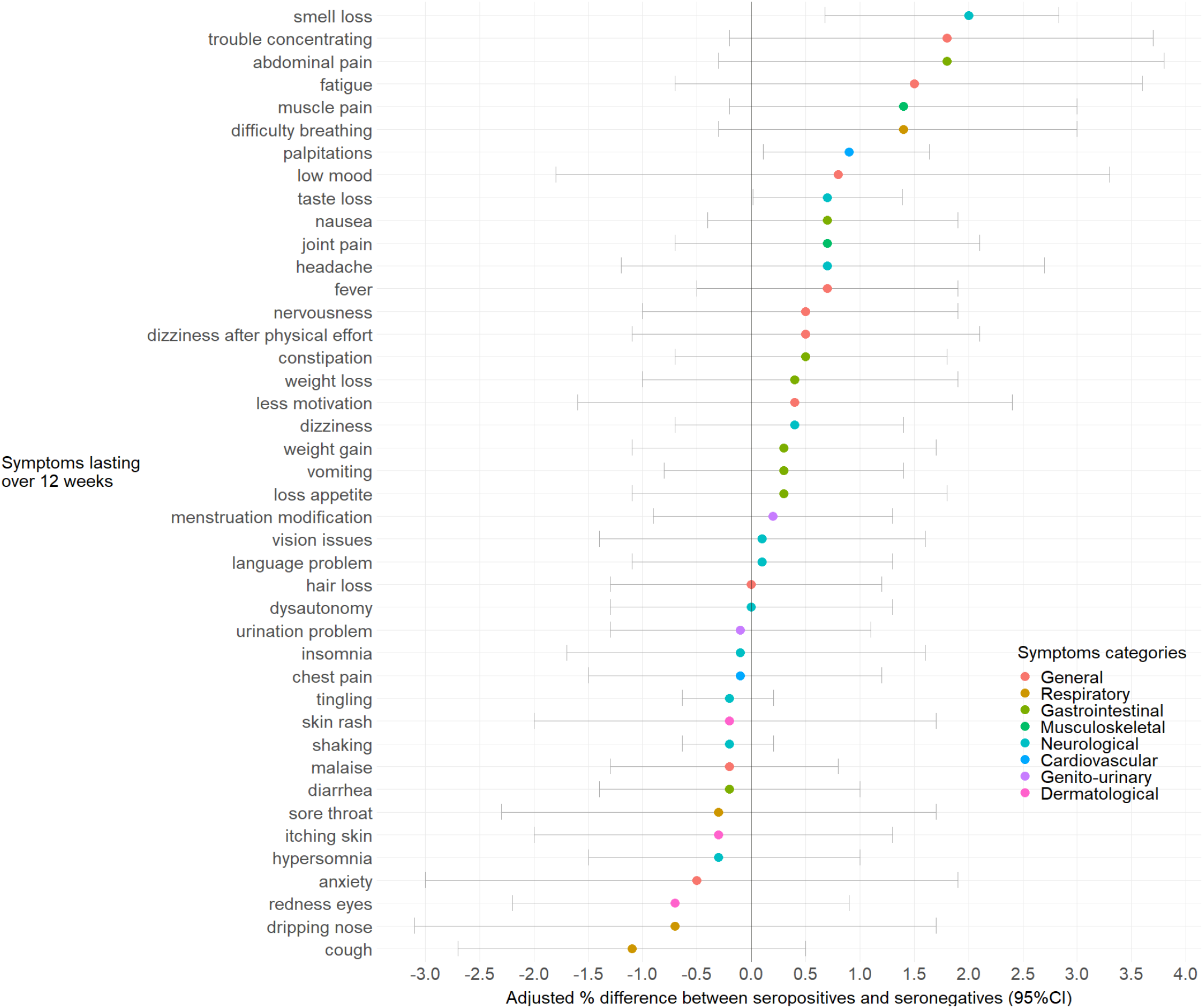
Sex- and age- adjusted difference (%) of symptoms lasting over 12 weeks between seropositives and seronegatives

### Adjusted prevalence of persistent symptoms overall and stratified by age

The adjusted prevalence of persistent symptoms among seropositive and seronegative children was 9.1% (95%CI: 6.7;11.8) and 5.0% (95%CI: 3.0;7.1), respectively. The corresponding adjusted prevalence difference (ΔaPrev) was 4.1% (95%CI: 1.1;7.3). After stratification by age groups, the prevalence of post-COVID syndrome was higher among adolescents (8.3%, 95%CI: 3.5;13.5) than among younger children (0.0%, 95%CI: −5.2;5.2 among 6-11 years old and 4.2%; 95%CI: −4.4;13.3 among 0-5 years old) (Table 2), in which no difference was observe.

**Table 2:**
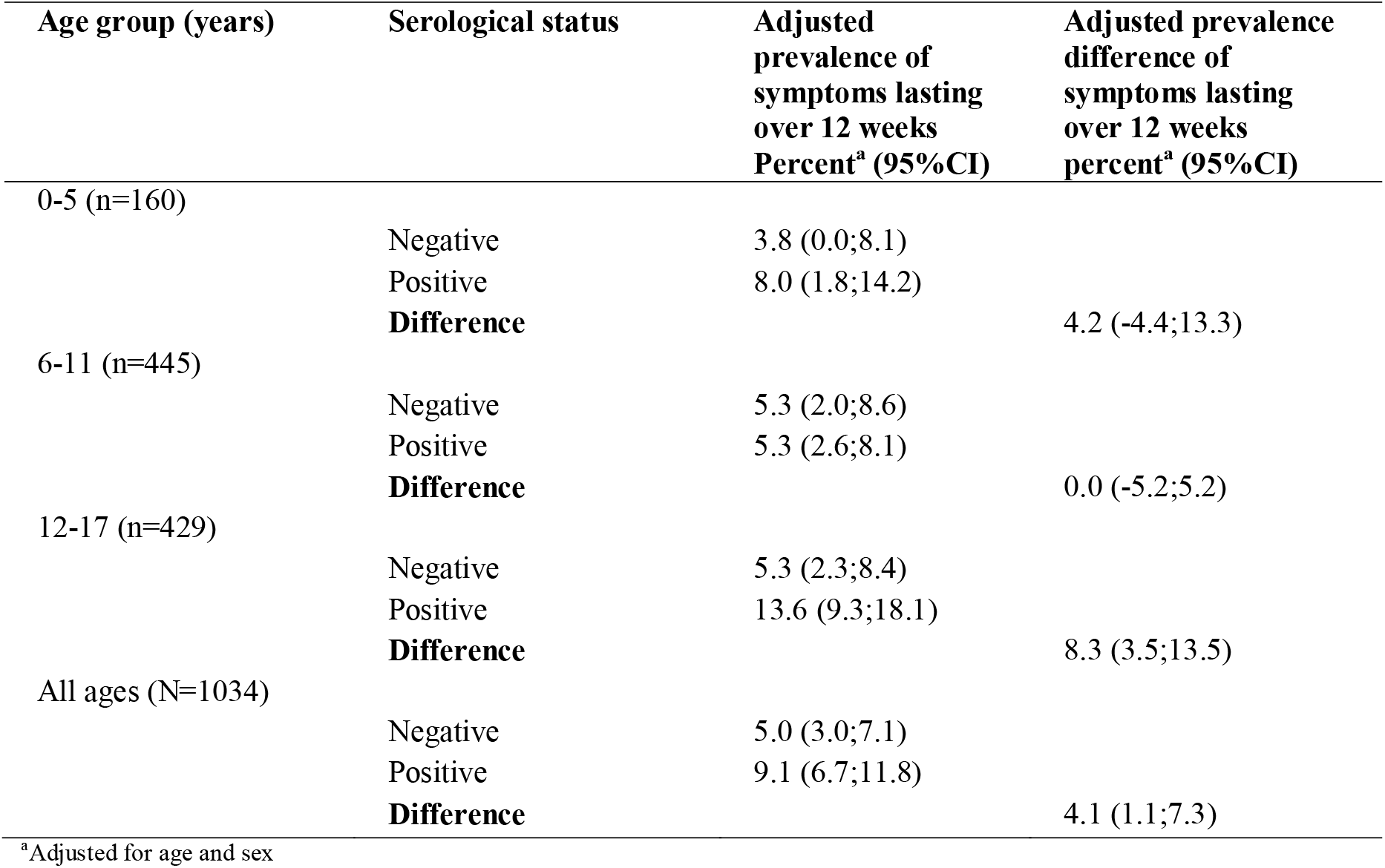
Sex and age-adjusted prevalence and prevalence difference of persistent symptoms

### Risk factors for persistent symptoms

Sex- and age-adjusted prevalence ratios were estimated for symptoms lasting over 12 weeks. Older age (aPR 1.1, 95%CI: 1.0;1.2, continuous variable), suffering from a chronic condition (aPR 3.6, 95%CI: 2.3;5.5) and living in a household with an average to poor financial situation (aPR 2.5, 95%CI: 1.4;4.6) were risk factors for experiencing persistent symptoms. Sex was not associated with long lasting symptoms (aPR 1.1, 95%CI: 0.8;1.6).

In a sub-analysis restricted to seropositive participants (N=570), we also observed that the risk factors associated with persistent symptoms were older age (aPR 1.1, 95%CI: 1.0;1.2), suffering from a chronic condition (aPR 3.5, 95%CI: 2.0;6.1) and living in a household with an average to poor financial situation (aPR 3.0, 95%CI: 1.5;6.2). Lower level of parental education was related to a higher risk of persistent symptoms, although not statistically significant (Table 3).

**Table 3:** Adjusted prevalence ratio of symptoms lasting over 12 weeks

| Report of symptoms lasting over 12 weeks |  | Overall<sup>a</sup> | Within seropositives<sup>a</sup> |
| --- | --- | --- | --- |
|  |  | N=1034<sup>b</sup> | n= 570<sup>b</sup> |
|  |  | PR (95%CI) | PR (95%CI) |
| **Sex** | Female | 1.0 (ref) | 1.0 (ref) |
|  | Male | 1.1 (0.8;1.6) | 1.1 (0.7;1.8) |
| **Age (years)** |  | 1.1 (1.0;1.2)* | 1.1 (1.0;1.3)* |
| **Chronic condition** | No | 1.0 (ref) | 1.0 (ref) |
|  | Yes | 3.6 (2.3;5.5)**<sup>c</sup> | 3.5 (2.0;6.1)**<sup>c</sup> |
| **Parental education** | Tertiary | 1.0 (ref) | 1.0 (ref) |
|  | Secondary | 1.2 (0.7;2.0)<sup>c</sup> | 1.3 (0.6;2.5)<sup>c</sup> |
|  | Primary | 1.9 (0.8;4.7)<sup>c</sup> | 1.7 (0.5;5.2)<sup>c</sup> |
| **Financial situation** | High | 1.0 (ref) | 1.0 (ref) |
|  | Average to poor | 2.5 (1.4;4.6)<sup>*c</sup> | 3.0 (1.5;6.2)<sup>*c</sup> |

<sup>a</sup>Prevalence ratio and 95% confidence interval are from Poisson regression with random effect on the household using the GLMMadaptive package in R and are adjusted for age, sex, or both according to independent variable.
<sup>b</sup>Complete case analysis. For each model presented, missing data in the covariates were excluded.
<sup>c</sup>Adjusting for age and sex
\*indicates p-value <0.05
\*\*indicates p-value <0.01

Based on the adjusted difference of frequency (%) of chronic health conditions within children experiencing 12 weeks persistent symptoms (n=40), we observed a significant positive difference between seropositives and seronegatives only with asthma (Table S3).

## Discussion

Our results showed that a significant proportion of children and adolescents suffered from persistent symptoms compatible with post-COVID syndrome. The overall estimated prevalence in the pediatric population was about 4%. Stratifying per age group, only adolescents displayed a substantial risk of having post-COVID symptoms. Risk factors for post-COVID syndrome were older age, having a lower socioeconomic status and suffering from chronic health conditions, especially asthma.

We defined post-COVID syndrome in children based on the recent definition published by Stephenson et al in March 2022 (2). The estimated overall prevalence of post-COVID syndrome in our sample is high, yet lower than in other studies (3). There might be multiple reasons. First, designs, samples and characteristics of post-COVID varied considerably across studies. Second, our data included mildly symptomatic and asymptomatic children and, therefore, a greater number of infected children than when relying on confirmed infection alone as testing was never systematic in children. Third, we worked with a population-based sample rather than clinical registries. Finally, a proportion of seropositive children in our sample might have been infected too recently making it impossible to studied long term persistent symptoms.

Our data showed a meaningful difference in prevalence between children older than 12 years and younger individuals, in agreement with the study by Stephenson et al (4). We could expect this proportion to be higher in reality as parents might not be aware of all of their children’s symptoms; adolescents may be less open about their issues since they are in a period of their life seeking more independence.

The prevalence difference between seronegatives and seropositives (∼8%) in adolescents is worth investigating as it potentially represents a very high absolute number, although lower than in adults (16). Post-COVID syndrome might worsen adolescents’ lives, already negatively impacted by the general pandemic environment (17,18) and could exacerbate long-term negative consequences on health, social and academic outcomes. On the contrary, we did not observe significant differences between seronegative and seropositive children younger than 12 years (19).

The pandemic-related difficulties to meet psychological and physical developmental needs may have been exacerbated by persistent symptoms, further preventing children from going about their daily activities and worsening their mental health. Most declared persistent symptoms among seropositives were smell loss, trouble concentrating, abdominal pain, fatigue, muscle pain, breathing difficulties and palpitations, in agreement with a recent literature review (3). The severity of symptoms reported by parents was, on average, slightly higher in seropositive children, suggesting that persistent symptoms following a SARS-CoV-2 infection might be more severe and represent a more substantial burden on their daily life (19). Sher et al. underlined that depressive symptoms appear to be common in patients with post-COVID syndrome. Long-term medical and psychological consequences of post-COVID syndrome could drastically worsen mental health and even increase suicidal ideation and behaviors (20). Similar outcomes could be expected in the pediatric population, especially in adolescents. Post-COVID condition should not be underestimated in children and calls for appropriate medical strategies including reinforcing vaccination strategies (21).

In an analysis restricted to seropositives, children from households with lower socioeconomic status were more likely to experience post-COVID syndrome. This is in line with the growing literature on health inequalities of the COVID-19 pandemic in terms of incidence, testing and severity of the infection (22). Those inequalities could be explained by differential exposure to the virus, greater susceptibility to infection, stronger comorbidities in vulnerable groups associated with severe outcomes, and disparities in healthcare. Similar mechanisms could explain the greater exposure of vulnerable children to the post-COVID syndrome. Also, it is reasonable to highlight that COVID-19 vaccination uptake was more frequent among individuals with higher socioeconomic status and is considered as the major protection against an acute or severe SARS-CoV-2 infection, hence developing a post-COVID syndrome (23).

In our sample, post-COVID seems more prevalent among children with chronic disease, especially asthma, although causality could be bi-directional. In general, asthmatic children have a higher risk of respiratory infections. However, the association between asthma and SARS-CoV-2 infection remains unclear in the pediatric population, as many large-scale ecological studies presented reduced pediatric asthma during the pandemic likely due to physical distancing, masks, and perhaps decreases in air pollution. On the other hand, other research focused on the individual level and presented asthma as a risk factor for hospitalization in children with COVID-19, but not for worse COVID-19 outcomes (24,25).

The major strength of our study is the population-based design with a wide age range, covering from babies to adolescents. Only very few studies on post-COVID syndrome rely on random population samples and include children of this age range (3). We identified previous SARS-CoV-2 infections relying on serological data, which also detects previous asymptomatic and mild infection. Unlike studies relying on test-confirmed infections, which suffer from selection bias, relying on serological assessment yields a better estimation of the proportion of the infected population.

However, the use of serological data also poses challenges. In particular, it is impossible to determine the exact date of infection only relying on this data (4). Our estimates are therefore based on a comparison of children with and without anti-SARS-CoV-2 antibodies, using a control group to avoid overestimating the prevalence of post-COVID symptoms. Once again, this highlights the importance of a control group and the complexity of identifying the post-COVID syndrome methodologically and clinically. Furthermore, relying on serological tests prevents the bias of parents over-reporting persistent symptoms when knowing their child(ren) have been infected (26).

Our study also has several limitations. Data were parent-reported and their answers could be influenced by their own experience or the household environment. Also, they might not be aware of some of their children’s symptoms, particularly for adolescents. Furthermore, as the time of infection was unknown in many cases, we are not able to study the incident risk of post-COVID, for which another study design including the time of the infection and a follow-up would be necessary. Moreover, our recruitment took place from December 2021, until February 2022, a period during which many children got infected (Omicron wave). Some children might suffer from long-lasting symptoms but were not identified as such in our analysis as they did not reach the 12-week threshold which could induce an underestimation of the post-COVID prevalence. It would be interesting to study persistent symptoms in children and adolescents who were more recently affected with the Omicron variant as less virulent variants could potentially lead to lower prevalence of post-COVID. Finally, participants seemed to come from a higher socioeconomic background than the Geneva population, therefore we might be underestimating the prevalence of persistent symptoms as socioeconomically disadvantaged individuals/households appear to be more likely to experience post-COVID.

## Conclusion

A significant proportion of children experienced post-COVID symptoms lasting over 12 weeks after infection with an estimated prevalence of 4% overall and 8% in adolescents. Older age, having a chronic condition and living on a household with lower socioeconomic conditions were identified as risk factors for the post-COVID syndrome. Our understanding of post-COVID syndrome will likely evolve as scientific evidence grows. Nevertheless, it is fundamental to rapidly implement effective primary care management, including early screening and detection, and health promotion to assist children and adolescents suffering from this syndrome who might experience long term physical and mental adverse consequences.

## Data Availability

Our data are accessible to researchers upon reasonable request for data sharing to the corresponding author.

## List of abbreviations

ΔaPrev: adjusted Prevalence Difference
aPrev: adjusted Prevalence
aPR: adjusted Prevalence Ratio
CI: Confidence interval
COVID-19: Coronavirus disease 2019
SARS-CoV-2: Severe acute respiratory syndrome coronavirus 2

## Acknowledgments

We are grateful to the staff of the Unit of Population Epidemiology of the HUG Division of Primary Care Medicine as well as to all participants whose contributions were invaluable to the study.

We also acknowledge all the members of the SEROCoV-KIDS study group: Deborah Amrein, Andrew S Azman, Antoine Bal, Michael Balavoine, Rémy P Barbe, Hélène Baysson, Julie Berthelot, Patrick Bleich, Livia Boehm, Gaëlle Bryand, Viola Bucolli, Prune Collombet, Alain Cudet, Vladimir Davidovic, Carlos de Mestral Vargas, Paola D’Ippolito, Richard Dubos, Roxane Dumont, Isabella Eckerle, Marion Favier, Nacira El Merjani, Natalie Francioli, Clément Graindorge, Idris Guessous, Séverine Harnal, Samia Hurst, Laurent Kaiser, Omar Kherad, Julien Lamour, Pierre Lescuyer, Arnaud G. L’Huillier, Andrea Jutta Loizeau, Elsa Lorthe, Chantal Martinez, Stéphanie Mermet, Mayssam Nehme, Natacha Noël, Francesco Pennacchio, Javier Perez-Saez, Anne Perrin, Didier Pittet, Jane Portier, Klara M Posfay-Barbe, Géraldine Poulain, Caroline Pugin, Nick Pullen, Viviane Richard, Frederic Rinaldi, Jessica Rizzo, Deborah Rochat, Cyril Sahyoun, Irine Sakvarelidze, Khadija Samir, Hugo Alejandro Santa Ramirez, Stephanie Schrempft, Claire Semaani, Silvia Stringhini, Stéphanie Testini, Yvain Tisserand, Deborah Urrutia Rivas, Charlotte Verolet, Jennifer Villers, Guillemette Violot, Nicolas Vuilleumier, Sabine Yerly, María-Eugenia Zaballa, Christina Zavlanou

## Ethics and dissemination

The SEROCoV-KIDS study was approved by the Cantonal Research Ethics Commission of Geneva, Switzerland (ID 2021-01973).

