## Supplementary material for "Post-COVID syndrome prevalence and risk factors in children and adolescents: A population-based serological study": S1

**A complete list of non-author contributors appears in the Acknowledgments.

**Table S1:** Descriptive statistics stratified by age group and serological status

|  | 0-5 ans^1^ | | 6-11 ans^1^ | | 12-17 ans^1^ | |
| --- | --- | --- | --- | --- | --- | --- |
|  | Negative,  N = 80 | Positive,  N = 80 | Negative,  N = 178^1^ | Positive,  N = 267 | Negativ,  N = 206 | Positive,  N = 223 |
| **Sex** |  |  |  |  |  |  |
| Female | 46 (57%) | 36 (45%) | 94 (53%) | 123 (46%) | 112 (54%) | 117 (52%) |
| Male | 34 (42%) | 44 (55%) | 84 (47%) | 143 (54%) | 94 (46%) | 106 (48%) |
| Other | 0 (0%) | 0 (0%) | 0 (0%) | 1 (0%) | 0 (0%) | 0 (0%) |
| **Chronic condition**^4^ | 9 (11%) | 8 (10%) | 38 (21%) | 37 (14%) | 73 (35%) | 76 (34%) |
| **Confirmed SARS-CoV-2 infection**^5^ | 3 (4%) | 16 (20%)** | 12 (7%) | 103 (39%)** | 10 (5%) | 109 (49%)** |
| **Confirmed SARS-CoV-2 symptomatic infection** | 3 (4%) | 14 (18%)* | 9 (5%) | 79 (30%)** | 9 (4%) | 84 (38%)** |
| **Vaccination Status**^7^ |  |  |  |  |  |  |
| No | 47 (100%) | 32 (100%) | 116 (99%) | 134 (100%) | 48 (36%) | 55 (48%) |
| Yes, 1 dose |  |  | 1 (1%) | 0 (0%) | 3 (2%) | 27 (23%) |
| Yes, 2 doses |  |  |  |  | 83 (62%) | 33 (29%) |
| **Persistent symptoms** |  |  |  |  |  |  |
| **Symptoms lasting over 4 weeks** | 14 (18%) | 11 (14%) | 31 (17%) | 36 (13%) | 28 (14%) | 52 (23%)* |
| Symptoms lasting 4 to 6 weeks | 5 (6%) | 4 (5%) | 15 (9%) | 14 (5%) | 7 (3%) | 12 (5%) |
| Symptoms lasting 6 to 8 weeks | 2 (2%) | 0 (0%) | 4 (2%) | 4 (2%) | 7 (3%) | 10 (4%) |
| Symptoms lasting 8 to 12 weeks | 4 (5%) | 0 (0%) | 2 (1%) | 2 (1%) | 2 (1%) | 2 (1%) |
| Symptoms lasting over 12 weeks | 3 (4%) | 7 (9%) | 10 (6%) | 16 (6%) | 12 (5%) | 31 (14%)* |

^1^Fisher's exact test or Pearson's Chi-squared test

*indicates p-value <0.05

**indicates p-value <0.01

**Table S2:** Characteristics of seropositive children with symptoms lasting over 12 weeks.

| Characteristic | n = 54 |
| --- | --- |
| **Sex** |  |
| Female | 25 (46%) |
| Male | 29 (54%) |
| Other | 0 (0%) |
| **Age group (years)** |  |
| 0-5 | 7 (13%) |
| 6-11 | 16 (30%) |
| 12-17 | 31 (57%) |
| **Confirmed SARS-CoV-2 infection^1^** | 30 (56%) |
| **Confirmed SARS-CoV-2 symptomatic infection^1^** | 26 (48%) |
| **Persistent symptoms declared after infection** | 17/30 (57%) |
| ^1^Diagnosed COVID-19 with a positive test (PCR, antigen test) | |

**Table S3:** Chronic condition among children who experienced symptoms lasting over 12 weeks

| **Characteristics** | Overall,  N = 40 | Seronegative,  n = 13 | Seropositive,  n = 27 | Sex and age-adjusted difference^a^  (Percent, 95%CI) |
| --- | --- | --- | --- | --- |
| **Asthma** | 10 (25%) | 1 (8%) | 9 (33%) | 25.3% (2.7;48.5)* |
| **Migraine** | 7 (18%) | 2 (15%) | 5 (19%) | 4.0% (-21.0;27.6) |
| **Obesity** | 3 (8%) | 1 (8%) | 2 (7%) | 0.3 % (-17.8;17.2) |
| **Osteo-articular**^b^ | 5 (12%) | 2 (15%) | 3 (11%) | -4.0 % (27.2;18.6) |
| **Dermatological**^c^ **condition** | 19 (20%) | 10 (31%) | 9 (15%) | -16.0% (-44.4;12.4) |

*indicates p-value <0.05

^a^Adjusting for age and sex

^b^Disease or malformation of the skeleton, joints, or muscles (e.g. kypho-scoliosis, scoliosis, lordosis, hip dysplasia, tendon rupture without trauma)

^c^Eczema, psoriasis,etc.
